# COVID-19 pandemic brings a sedentary lifestyle: a cross-sectional and longitudinal study

**DOI:** 10.1101/2020.05.22.20110825

**Authors:** Chen Zheng, Wendy Yajun Huang, Sinead Sheridan, Cindy Hui-Ping Sit, Xiang-Ke Chen, Stephen Heung-Sang Wong

## Abstract

**Objectives:** The coronavirus disease 2019 (COVID-19) pandemic continues to pose profound challenges on society. Governments around the world have managed to mitigate its spread through strategies including social distancing; however, this may result in the adoption of sedentary lifestyle. This study aimed to investigate: 1) physical activity (PA) levels, sedentary behavior (SB) and sleep among young adults during COVID-19 epidemic, and 2) the change in these behaviors before and during the pandemic.

**Methods:** A total of 631 young adults (38.8% males) participated in the cross-sectional study and completed an online survey that included five components: general information, COVID-19 related issues, PA, SB, and sleep. For longitudinal study, PA, SB, and sleep data collected from 70 participants before and during COVID-19 pandemic were analyzed.

**Results:** Participants reported engaging in low PA, high SB and long sleep duration during COVID-19 pandemic. Females had greater concern for COVID-19 related issues and engaged in more prevention strategies than males. Moreover, a significant decline in PA while increase in both times spent in SB and sleep were determined after COVID-19 outbreak.

**Conclusion:** The results of this study demonstrated a sedentary lifestyle in young adults in responses to social distancing during the COVID-19 pandemic, which will assist health policy makers and practitioners in the development of population specific health education and behavior interventions during this pandemic and for other future events.

*What are the new findings?:*

- This is the first study to investigate lifestyle behaviors in young adults during the COVID-19 pandemic and the changes in these behaviors after its outbreak.
- Low physical activity (PA) level, high sedentary behavior (SB) and long sleep duration were found in young adults during the COVID-19 pandemic.
- All types of PA and both time spent in SB and sleep significantly decreased and increased after COVID-19 outbreak, respectively.

*How might it impact on clinical practice in the future?:* The current study provided evidences that young adults engaged in sedentary lifestyle during the COVID-19 pandemic, which may assist in the development and provision of appropriate and tailored health education and behavior interventions during and after this or other future global pandemics.

## INTRODUCTION

Since the first outbreak of the coronavirus disease 2019 (COVID-19) in Wuhan, China, in early December 2019, [1] the disease has rapidly spread across the world, with the first confirmed case in Hong Kong reported in late January 2020. [2] Since then, unprecedented efforts have been made by the Governments around the world to slow the incidence of infection. The efforts made by Hong Kong Government, for instance, included border entry restrictions, quarantine and isolation of cases and contacts, and closure of schools resulting in major disruptions to daily routines.[3] An escalation in the number of cases in Hong Kong in late March 2020 further fueled the Government to enforce stricter measures including the closure of leisure facilities and cultural facilities,[4] and the continued delivery of courses to students via online platforms for the remainder of the academic term.

While these measures are highly commendable and critical to mitigate the spread of COVID-19, they may result in inducing unhealthy behaviors like sedentary lifestyle, with most individuals adhering to social distancing by working or studying from home or in other cases, self-isolating under strict quarantine. Of particular concern, is the potential deleterious effects of reduced physical activity (PA) and increased sedentary behaviors (SB) that coincide with social distancing on both physical and mental health outcomes.

Under normal circumstances, a sedentary lifestyle including physical inactivity and prolonged SB have been previously identified as problematic among adults. [5] Social distancing, including closure of schools and home confinement, has been shown to result in less PA, prolonged SB and experience poor sleep quality.[6] However, the impacts of social distancing during COVD-19 pandemic on lifestyle behaviors is currently unknown due to the lacking of evidences. Thus, this study aimed to investigate: 1) PA levels, time spent in SB, and sleep in Hong Kong young adults during the COVID-19 pandemic; and 2) the changes in these lifestyle behaviors after COVID-19 outbreak.

## METHODS

### Study design and participants

The study design included both cross-sectional and longitudinal trials. For recruitment for cross-sectional trial, information was advertised online and through word of mouth. Inclusion criteria for study participation included: 1) adults aged 18-35 years old and 2) living in Hong Kong for the past two months. Participants completed an online survey supported by Google form (Google LLC, U.S.A.) which included five components: general information (e.g., age, sex, body weight, and height), COVID-19 related issues, PA, SB, and sleep. Participant body mass index (BMI) was subsequently calculated as weight in kilograms divided by height in meters square. The online survey was conducted between April 15, 2020 and April 26, 2020, with a total of 631 respondents. For the longitudinal trial, 70 young adults from our previous research study conducted in 2019 were invited to report their lifestyle behaviors during the pandemic and further compared with their data of lifestyle behaviors before COVID-19 outbreak. Of those participants, 60 completed all three questionnaires used to assess their PA, SB, and sleep while 10 participants only completed the PA questionnaire.

### Physical activity

The International Physical Activity Questionnaires (IPAQ) was used to assess the PA level in participants. The validity and reliability of the short version IPAQ has been tested in 12 countries,[7] which has been shown to be suitable for population surveillance and large-scale studies. In the present study, three of the seven items were taken from the questionnaire to obtain information on engagement of participants in vigorous PA (VPA), moderate PA (MPA), and walking. The MET-minutes per week (MET.min/week) were calculated using the following formula: intensity (MET) x duration x frequency. In addition, to assess the impact of COVID-19 on PA, one more question was asked, “*How has your physical activity levels been since the onset of the COVID-19 pandemic?* (e.g., increase, no change, and decrease)”.

### Sedentary behavior

SB was measured using the Sedentary Behavior Questionnaire (SBQ) in participants, which has been previously validated in adults. [8] Intra-class correlation coefficients for all nine items and total scale were acceptable (range = 0.51 - 0.93).[8] A total of nine SB (TV/DVD, computer/video games, sit listening to music, sit talk on telephone, doing computer/paper work, reading books, playing musical instrument, doing art and crafts, and sitting for transport) were selected for this questionnaire. All items were assessed for a usual weekday and weekend day for the past month with nine options: none, ≤15 min, 30 min, 1 h, 2 h, 3 h, 4 h, 5 h, and ≥6 h. Based on previous methodology published, the time spent on each behavior was converted into hours (e.g., a response of 15 min was recorded as 0.25 h).[8] To obtain daily estimates, each item of weekday hours were multiplied by 5 and weekend hours were multiplied by 2, and these were then divided by 7. The daily SB was assessed by IPAQ, which a separate question asking for sitting time.[8]

### Sleep

The most commonly subjective sleep scale, the Pittsburgh Sleep Quality Index (PSQI), was used to assess both sleep quality and sleep duration in participants. The PSQI is a validated 19-item, self-reported questionnaire, which is categorized into seven sleep quality components (subjective sleep quality, sleep latency, sleep duration, habitual sleep efficiency, sleep disturbances, use of sleeping medication, and daytime dysfunction).[9] The final score of the seven sleep components ranged from 0 to 21 points. Final scores of 5 or < 5 points are classified as having “good sleep quality”, and > 5 points is classified as “poor sleep quality”.[9] The sleep duration was calculated from participants reported bed and wake-up times. Besides, to assess the impact of COVID-19 on sleep quality, one more question was asked, “*How was your sleep since the onset of the COVID-19 pandemic?* (e.g., better than usual, the same as usual, and worse than usual)”.

### COVID-19 related issues

Participants were also asked the following five questions related to COVID-19; (1) *“Please identify your main source of information regarding the COVID-19 pandemic:* Newspapers or Television, Government websites, Work colleagues/friends, Facebook/Twitter/Instagram/YouTube”; (2) *“Have you ever been home quarantined or stayed in a quarantine center for compulsory quarantine?* Yes, No or Prefer not to say”; (3) “I *am concerned about contracting COVID-19 myself”;* (4) *“I am concerned about other family members or friends contracting COVID-19”*. Answer from participants for question (3) and (4) using one of the following five options: “not at all concerned, slightly concerned, somewhat concerned, moderately concerned, extremely concerned”. (5) *“How often do you practice these prevention strategies against the spread of COVID-19?”* For this question, three of the most common and effective prevention strategies methods were chosen including “regular hand-washing with soap, wearing a face mask, and avoiding restaurants/gyms/shops”. All three items were answered using the following five options: “always, often, sometimes, rarely, and never”.

### Statistical analysis

Three international guidelines for PA, SB and sleep for adults for health were applied for data analysis: (1) achievement of at least 150 min of moderate-intensity aerobic PA or at least 75 min of vigorous-intensity aerobic PA throughout the week,[10] (2) engagement in < 9 h of SB per day for adults,[11] (3) score of sleep quality < 5 with sleep duration between 7 and 9 h.[12] Descriptive information (all and stratified by sex), including participant characteristics, COVID-19 related issues, and participants’ daily behaviors, were summarized and reported as means ± standard deviation (SD) or median (interquartile range) for continuous variables and as proportions of participants for categorical variables. Independent samples t tests and Chi Square tests were used to assess the difference between males and females for continuous variables and categorical variables, respectively. The change in participants’ daily behavior (e.g., PA, SB and sleep) were determined using paired sample t tests and shown as means ± SD. All statistical tests were performed using SPSS for Windows, version 24 (IBM Corp., Armonk, N.Y., USA).

### Patient and public involvement

Patients were not involved in this research to comment on the study design or interpret the results. Patients were not invited to contribute in the writing or editing of the manuscript.

## RESULTS

### Descriptive statistics of participant and COVID-19 related issues

A total of 631 participants (mean age ± SD, 21.1 ± 2.9 years, 38.8% males) was included in data analysis. The descriptive statistics for the characteristics of participants were shown in Table 1. It showed the COVID-19 related issues in Table 2, including main sources of information, the concern of contracting COVID-19, and prevention strategies. Generally, females had greater concern for contracting COVID-19 themselves and for their family members contracting COVID-19. Therefore, females engaged more in COVID-19 prevention strategies such as wearing a face mask and avoiding restaurants, gyms, and shops compared with males.

**Table 1.** Participant characteristics for cross-sectional study, and stratified by sex.

| | Mean ± SD | | |
| --- | --- | --- | --- |
|  | All (n=631) | Males (n=245) | Females (n=386) |
| Age (years)* | 21.1 ± 2.9 | 21.5 ± 3.2 | 20.9 ± 2.5 |
| Height (cm)* | 165.6 ± 8.3 | 173.2 ± 6.1 | 160.8 ± 5.5 |
| Weight (kg)* | 57.0 ± 10.1 | 64.3 ± 10.0 | 52.4 ± 7.0 |
| BMI (kg/m<sup>2</sup>)* | 20.7 ± 2.6 | 21.4 ± 2.9 | 20.3 ± 2.4 |

BMI: body mass index; SD: standard deviation
\*Gender differences, $p < 0.05$

**Table 2.** COVID-19 related issues for cross-sectional study and stratified by sex.

|  | % |  |  |
| --- | --- | --- | --- |
|  | All (n=631) | Males (n=245) | Females (n=386) |
| Main sources of information |  |  |  |
| Newspapers or TV | 37.4 | 33.5 | 39.9 |
| Government websites | 2.4 | 1.2 | 3.1 |
| Work colleagues/friends | 1.0 | 0.8 | 1.0 |
| Facebook/Twitter/Instagram/YouTube | 59.3 | 64.5 | 56.0 |
| Home quarantine/compulsory quarantine |  |  |  |
| Yes | 7.1 | 5.7 | 8.0 |
| No | 92.1 | 93.1 | 91.5 |
| Prefer not to say | 0.8 | 1.2 | 0.5 |
| Contracting COVID-19 myself** |  |  |  |
| Not at all concerned | 3.2 | 5.7 | 1.6 |
| Slightly concerned | 13.6 | 15.5 | 12.4 |
| Somewhat concerned | 22.5 | 25.3 | 20.7 |
| Moderately concerned | 43.1 | 37.6 | 46.6 |
| Extremely concerned | 17.6 | 15.9 | 18.7 |
| Family members contracting COVID-19* |  |  |  |
| Not at all concerned | 1.4 | 2.4 | 0.8 |
| Slightly concerned | 9.0 | 12.2 | 7.0 |
| Somewhat concerned | 20.6 | 23.3 | 18.9 |
| Moderately concerned | 43.4 | 38.4 | 46.6 |
| Extremely concerned | 25.5 | 23.7 | 26.7 |
| Prevention strategies |  |  |  |
| Hand-washing with soap |  |  |  |
| Always | 55.6 | 49.8 | 59.3 |
| Often | 32.5 | 35.5 | 30.6 |
| Sometimes | 9.4 | 11.0 | 8.3 |
| Rarely | 2.2 | 2.9 | 1.8 |
| Never | 0.3 | 0.8 | 0.0 |
| Wearing a face mask* |  |  |  |
| Always | 88.9 | 84.9 | 91.5 |
| Often | 8.1 | 9.4 | 7.3 |
| Sometimes | 2.1 | 3.7 | 1.0 |
| Rarely | 0.5 | 0.8 | 0.3 |
| Never | 0.5 | 1.2 | 0.0 |
| Avoiding restaurants/gyms/shops* |  |  |  |
| Always | 35.0 | 28.6 | 39.1 |
| Often | 34.2 | 34.7 | 33.9 |
| Sometimes | 24.4 | 27.3 | 22.5 |
| Rarely | 5.5 | 8.2 | 3.9 |
| Never | 0.8 | 1.2 | 0.5 |

\*Gender differences, $p < 0.05$ ; \*\* $p < 0.01$

### Sedentary lifestyle during COVID-19 pandemic

Based on BMI and cut-off point of overweight and obesity, 12.2% and 23.3% of the males and females were overweight, respectively. Lifestyle behaviors (e.g., PA, SB, and sleep) are presented in Table 3, where 30.0% of participants met the PA guideline, and greater than half of the participants (57.8%) did not engage in any VPA during the COVID-19 pandemic. 70% of participants reported that their PA level decreased since the onset of the COVID-19 pandemic. Compared with males, engagement in computer/video games was lower while engagement in computer/paper work and arts and crafts was higher in females. Despite females had a significantly longer mean sleep duration than males (8.7 ± 1.2 vs. 8.5 ± 1.2, p<0.05), more male participants met the sleep guideline than females (46.5 vs 38.1, p<0.05).

**Table 3.** Participants’ lifestyle behaviors for cross-sectional study and stratified by sex.

| | Median (IQR) or Mean ± SD or % | | |
| --- | --- | --- | --- |
|  | All (n=631) | Males (n=245) | Females (n=386) |
| PA |  |  |  |
| Walking (min/day) | 17.1 (28.6) | 17.1 (27.1) | 12.85 (28.6) |
| Moderate activity (min/day) | 2.9 (11.4) | 2.85 (12.9) | 2.85 (10.4) |
| Vigorous activity (min/day)* | 0.0 (8.6) | 0.0 (10.0) | 0.0 (8.6) |
| Total energy expenditure (MET.min/week)** | 792.0 (1398.0) | 864.0 (1836.0) | 792.0 (1230.0) |
| Change of PA level |  |  |  |
| Increase | 16.5 | 12.2 | 19.2 |
| No change | 11.3 | 10.6 | 11.7 |
| Decrease | 72.3 | 77.1 | 69.2 |
| Meet PA guideline<sup>1</sup> | 29.6 | 30.2 | 29.3 |
| SB |  |  |  |
| TV/DVD (h/day) | 1.7 ± 1.5 | 1.6 ± 1.5 | 1.8 ± 1.5 |
| Computer/video games (h/day)** | 1.4 ± 1.6 | 1.9 ± 1.8 | 1.1 ± 1.4 |
| Sit listen to music (h/day) | 1.1 ± 1.3 | 1.1 ± 1.4 | 1.1 ± 1.3 |
| Sit talk on telephone (h/day) | 0.8 ± 1.2 | 0.9 ± 1.3 | 0.8 ± 1.2 |
| Computer/paper work (h/day)** | 3.1 ± 1.8 | 2.9 ± 1.8 | 3.3 ± 1.8 |
| Reading books (h/day) | 0.5 ± 0.9 | 0.5 ± 0.9 | 0.6 ± 0.9 |
| Play musical instrument (h/day) | 0.2 ± 0.5 | 0.2 ± 0.4 | 0.2 ± 0.6 |
| Doing arts and crafts (h/day)** | 0.2 ± 0.5 | 0.1 ± 0.4 | 0.2 ± 0.6 |
| Sitting for transport (h/day) | 0.4 ± 0.6 | 0.4 ± 0.7 | 0.4 ± 0.5 |
| Daily SB (h/day) | 9.4 ± 3.0 | 9.4 ± 3.2 | 9.5 ± 3.0 |
| Meet SB guideline<sup>2</sup> | 42.5 | 44.1 | 41.5 |
| Sleep |  |  |  |
| Sleep duration (h/day)* | 8.6 ± 1.2 | 8.5 ± 1.2 | 8.7 ± 1.2 |
| Sleep quality | 5.2 ± 2.5 | 4.9 ± 2.3 | 5.3 ± 2.6 |
| Meet sleep guideline<sup>3</sup>* | 41.4 | 46.5 | 38.1 |
| Change of sleep quality |  |  |  |
| Better than usual | 18.4 | 20.8 | 16.8 |
| The same as usual | 43.9 | 46.9 | 42.0 |
| Worse than usual | 37.7 | 32.2 | 41.2 |

PA: physical activity; SB: sedentary behavior; IQR: interquartile range; SD: standard deviation
<sup>1</sup>At least 150 min of moderate-intensity aerobic PA or at least 75 min of vigorous-intensity aerobic PA throughout the week
<sup>2</sup> Less than 9 h of SB per day
<sup>3</sup> The score of sleep quality < 5 with the sleep duration in 7-9 h
\*Gender differences, $p < 0.05$ ; \*\* $p < 0.01$

### Changes in lifestyle behaviors after COVID-19 outbreak

The changes in lifestyle behaviors in participants from the longitudinal trial was presented in Figure 1. After COVID-19 outbreak, all PA levels, including VPA (before vs. during COVID-19, 9.5 ± 12.5 vs. 6.0 ± 11.6, p<0.05), MPA (11.2 ± 16.0 vs. 5.5 ± 8.7, p<0.01), and walking (39.7 ± 30.7 vs. 19.8 ± 24.5, p<0.01) were significantly declined while both time spent in SB (7.8 ± 3.2 vs. 10.0 ± 3.2, p<0.01) and sleep duration (7.7 ± 1.0 vs. 8.4 ± 1.2, p<0.01) significantly increased. A lower percentage of participants met the guidelines for PA (50.0% vs. 20.0%), SB (54.3% vs. 40.0%), and sleep (67.1% vs. 57.1%) after COVID-19 outbreak. When considering the SB by types of activities, engagement in TV/DVD (0.9 ± 0.8 vs. 1.7 ± 1.4, p<0.01) and computer/paper work (2.2 ± 1.7 vs. 3.1 ± 2.0, p<0.01) was significantly higher and sitting time during transportation (0.7 ± 0.7 vs. 0.4 ± 0.6, p<0.01) was significantly lower during COVID-19 than before the epidemic.

**Figure 1.**
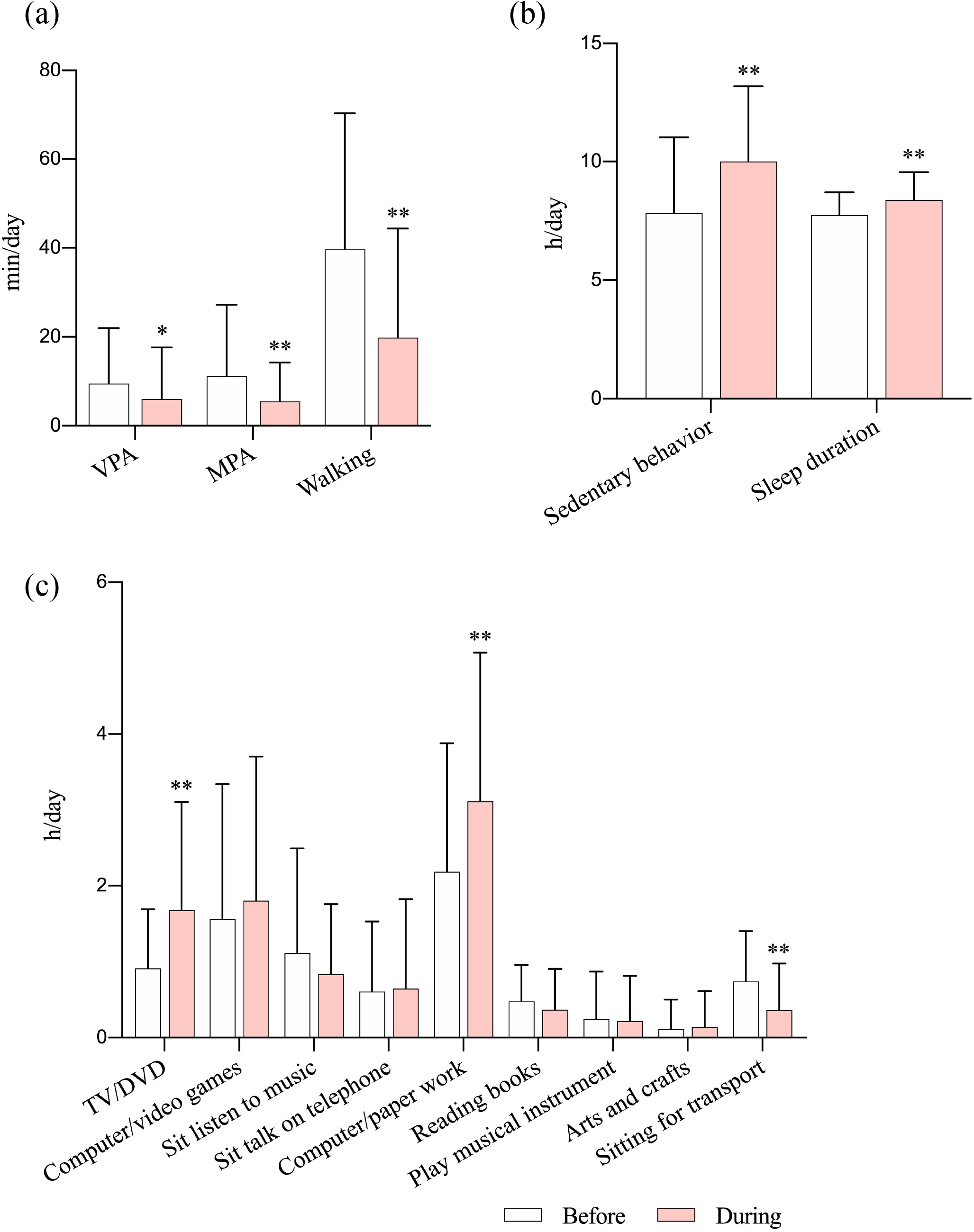
The changes in participants’ lifestyle behaviors. (a) Physical activity, (b) sedentary behavior and sleep duration, (c) sedentary behavior by types by activities MPA: moderate physical activity; VPA: vigorous physical activity *p<0.05, **p<0.01, compared with before.

## DISCUSSION

This is first study, to the best of our knowledge, to investigate PA levels, time spent in SB, and sleep in young adults during COVID-19 pandemic and the changes in these lifestyle behaviors after COVID-19 outbreak. The major findings of our study were that engagement in all PA behaviors significantly declined while both time spent in SB and sleep duration significantly increased after COVID-19 outbreak, which demonstrated a sedentary lifestyle during this pandemic.

### COVID-19 related issues

Individual behaviors of young adults have been changed in response to the threat posed by COVID-19. Based on our results, 90% of participants reported that they always wear a face mask when leaving home, and only 0.3% of participants reported never wearing a face mask, which may also contribute to the reduction in PA and exercise, as wearing masks may increase the difficulty in breathing during outdoor activities. A similar result was previously reported in that 99% of participants reported wearing face masks when leaving home.[3] In addition, 85% of participants reported that they always or often wash their hands with soap. When the severe acute respiratory syndrome (SARS) occurred in Hong Kong in 2003, the proportion of face mask use and washing hands among adults was 79% and 82% respectively. [13] Interestingly, females showed more concerns about contracting COVID-19 themselves or their family members contracting it, thus perform more prevention strategies that always wore a face mask and avoided restaurants/gyms/shops than males, which may lead to gender-difference in the reduced outdoor activities during the pandemic.

### Physical activity and sedentary behavior

In the present study, all types of PA (i.e., MPA, VPA and walking) significantly decreased after COVID-19 outbreak. Cross-sectional analysis also revealed the low volume of PA that participants engaged in during COVID-19 pandemic, with average 3min/day MPA and 17min/day walking. The low volume of PA undertaken by participants may be due to social distancing (e.g., cancellation of all team sports training and competitions and the closure of public leisure facilities and gymnasiums) and the concerns of threat posed by COVID-19 in Hong Kong. Furthermore, the limited living space in Hong Kong, which is smaller than in other Asian cities including Tokyo and Singapore, may further restrict the opportunities of young adults to exercise at home.[14]

The total time spent in SB during the waking day was significantly higher during COVID-19 than prior to the outbreak. Specifically, time spent in both TV/DVD and computer/paper work significantly increased while sitting time for transport decreased after COVID-19 outbreak. This may be partially explained by young adults engaging in social distancing by staying home and online teaching, which subsequently resulted in prolonged screen time, such as elevated time spent in watching TV, playing computer game and online teaching.[6]

Increased screen time found in this study has been previously shown to be accompanied by prolonged sedentary bouts without interruptions which have a more negative impact on health outcomes.[15] Interventions that increase PA while decrease SB in young adults during and after COVID-19 pandemic are warranted. For instance, simple home-based exercises, such as body resistance training or high-intensity interval training (HIIT), can be applied to this population. Appropriate health education and behavior interventions via online platforms can be also developed during this pandemic, as more than 70% of participants reported that their engagement in PA had significantly decreased during the COVID-19 pandemic.

### Sleep

The COVID-19 pandemic brings significant disruptions to daily routines. With a more flexible schedule due to the closure of school and companies, participants had significantly longer sleep duration during this pandemic. Of particular interest is that 37% of participants reported having poorer sleep quality during COVID-19 pandemic. Since poor sleep is highly correlated with stress,[16] worse sleep quality reported by participants may due to the stress induced by the threat of COVID-19. The effect of COVID-19 on sleep and mental health among young adults warrants further investigation.

### Limitations and strengths

One major strength of this study is that both cross-sectional and longitudinal analysis were applied in this study. Secondly, there was a relatively large sample size in the cross-sectional analysis. Accordingly, the sample was representative of the young adults in Hong Kong. Thirdly, three behaviors (PA, SB, and sleep), which occupy a large proportion of time in individuals over 24 hours were assessed in the current study.

The limitations of this study include the use of subjective measurements to assess PA, SB and sleep, which are associated with increased risk of bias. Though all the questionnaires used in this study have been previously validated, objective measurement, such as use of accelerometer, would be more accurate in assessing PA and SB in participants.

## CONCLUSION

Low PA levels, high time spent in SB and long sleep duration were identified in young adults during the COVID-19 pandemic, with less than half of the participants meeting any of the recommended guidelines for PA, SB or sleep. There was also a significant reduction in PA behaviors and a significant increase in SB and sleep duration of young adults after COVID-19 outbreak. These findings may have important public health implications and provide evidences for future intervention studies.

## Data Availability

All data relevant to the study are included in the article.

## Contributors

CZ, XC and SHW designed the study. CZ, WYH, and XC collected the data. CZ, WYH, and CHS analyzed the data. CZ and SS wrote the first draft. All authors contributed to the final draft.

## Acknowledgments

We would like to thank all participants for their efforts and participation in this study. We thank Ms. ZHANG Xiaoyuan and Mr. Waris WONGPIPIT from Department of Sports Science and Physical Education, Faculty of Education, The Chinese University of Hong Kong for their support during the data collection.

## Ethics approval

Survey and Behavioral Research Ethics, The Chinese University of Hong Kong (SBRE-19-599).

## Data availability statement

All data relevant to the study are included in the article.

## REFERENCE

1 Wu JT, Leung K, Leung GM. Articles Nowcasting and forecasting the potential domestic and international spread of the 2019-nCoV outbreak originating in Wuhan, China: a modelling study. Lancet 2020;395:689–97. doi:10.1016/S0140-6736(20)30260-9

2 news.gov.hk. Health declaration applies to XRL. 2020. https://www.news.gov.hk/eng/2020/01/20200123/20200123_203125_697.html?type=category&name=covid19&tl=t (accessed 3 May 2020).

3 Cowling BJ, Ali ST, Ng TWY, et al. Impact assessment of non-pharmaceutical interventions against COVID-19 and influenza in Hong Kong: an observational study. Lancet Public Heal 2020;2667:2020.03.12.20034660. doi:10.1101/2020.03.12.20034660

4 The Government of the Hong Kong Special Administrative Region Press Releases. Latest arrangements on LCSD public services. 2020. https://www.info.gov.hk/gia/general/202003/21/P2020032100740.htm?fontSize=1 (accessed 26 Apr 2020).

5 Hallal PC, Andersen LB, Bull FC, et al. Global physical activity levels: Surveillance progress, pitfalls, and prospects. Lancet 2012;380:247–57. doi:10.1016/S0140-6736(12)60646-1

6 Wang G, Zhang Y, Zhao J, et al. Mitigate the effects of home confinement on children during the COVID-19 outbreak. Lancet 2020;395:945–7. doi:10.1016/S0140-6736(20)30547-X

7 Craig CL, Marshall AL, Sjöström M, et al. International physical activity questionnaire: 12-Country reliability and validity. Med Sci Sports Exerc 2003;35:1381–95. doi:10.1249/01.MSS.0000078924.61453.FB

8 Rosenberg DE, Norman GJ, Wagner N, et al. Reliability and validity of the sedentary behavior questionnaire (SBQ) for adults. J Phys Act Heal 2010;7:697–705. doi:10.1123/jpah.7.6.697

9 Buysse, D. J., Reynolds, C. F., Monk, T. H., Berman, S. R., & Kupfer DJ. The Pittsburgh Sleep Quality Index: a new instrument for psychiatric practice and research. Psychiatry Res 1989;8:193–213.

10 World Health Organization. Physical Activity and Adults. 2020. https://www.who.int/dietphysicalactivity/factsheet_adults/en/ (accessed 29 Apr 2020).

11 Ku PW, Steptoe A, Liao Y, et al. A cut-off of daily sedentary time and all-cause mortality in adults: A meta-regression analysis involving more than 1 million participants. BMC Med 2018;16:1–9. doi:10.1186/s12916-018-1062-2

12 National Sleep Foundation. National Sleep Foundation Recommends New Sleep Times. 2015. https://www.sleepfoundation.org/press-release/national-sleep-foundation-recommends-new-sleep-times (accessed 29 Apr 2020).

13 Leung, G. M., Quah, S., Ho, L. M., Ho, S. Y., Hedley, A. J., Lee, H. P., & Lam TH. A tale of two cities: community psychobehavioral surveillance and related impact on outbreak control in Hong Kong and Singapore during the severe acute respiratory syndrome epidemic. Infect Control Hosp Epidemiol 2004;25:1033–41.

14 Our Hong Kong Foundation. Land and Housing | Policy Research & Advocacy Series. 2020. https://www.ourhkfoundation.org.hk/en/report/18/land/land-and-housing-policy-research-advocacy-series (accessed 30 Apr 2020).

15 Carson V, Wong SL, Winkler E, et al. Patterns of sedentary time and cardiometabolic risk among Canadian adults. Prev Med (Baltim) 2014;65:23–7. doi:10.1016/j.ypmed.2014.04.005

16 Hirotsu C, Tufik S, Andersen ML. Interactions between sleep, stress, and metabolism: From physiological to pathological conditions. Sleep Sci 2015;8:143–52. doi: 10.1016/j.slsci.2015.09.002

